# Functional genomics for curation of variants in telomere biology disorder associated genes, a systematic review

**DOI:** 10.1101/2022.07.04.22277240

**Authors:** Niles Nelson, Simone Feurstein, Aram Niaz, Jia Truong, Jessica K. Holien, Sionne Lucas, Kirsten Fairfax, Joanne Dickinson, Tracy M. Bryan

**Author notes:** Correspondence and requests for materials should be addressed to Niles Nelson, The Menzies Institute for Medical Research, The University of Tasmania, Hobart, Tasmania, Australia. equal last author (Joanne Dickinson, Tracy M. Bryan).

## Abstract

**Background:** Patients with an underlying telomere biology disorder (TBD) have variable clinical presentations and can be challenging to diagnose clinically. A genomic diagnosis for patients presenting with TBD is vital for optimal treatments. Unfortunately, many variants identified during diagnostic testing are variants of uncertain significance (VOUS). This complicates management decisions, delays treatment and risks non-uptake of a potentially curative therapies. Improved application of functional genomic evidence may reduce VOUS classifications.

**Methods:** We systematically searched the literature for published functional assays interrogating TBD gene variants. Where possible, established likely benign/benign and likely pathogenic/pathogenic variants were used to estimate the assay sensitivity, specificity, positive predictive value, negative predictive value and odds of pathogenicity.

**Results:** 3131 articles were screened and 152 met inclusion criteria. Sufficient data to enable a PS3/BS3 recommendation was available for *TERT* variants only. We recommend PS3 and BS3 can be applied at a moderate and supportive level respectively. PS3/BS3 application was limited by a lack of assay standardisation and limited inclusion of benign variants.

**Conclusions:** Further assay standardisation and assessment of benign variants is required for optimal use of the PS3/BS3 criterion for TBD gene variant classification.

## Introduction

### Telomere biology disorder

Renewal and regeneration of body tissues is essential throughout life. Cells are replaced by proliferation of stem cell populations located throughout the body. Due to RNA primer placement on the lagging strand, replication stops 50-100 base pairs (bp) prior to the end of the chromosome with each replication. To avoid erosion of essential coding and regulatory genomic DNA, each chromosome is capped with telomeres. Inherited defects in telomere maintenance pathways result in⍰telomere biology disorder (TBD), a disorder with diverse multi-organ phenotypes associated with the loss of stem cell populations^1-4^. Currently, germline loss of function variants in 14 genes (*ACD, CTC1, DKC1, NAF1, NHP2, NOP10, PARN, RTEL1, TERC, TERT, TINF2, STN1, WRAP53*, and *ZCCHC8*) are described as causative of TBD^3,5^. Individuals with TBD can present with pathologies including bone marrow failure, myelodysplasia, acute myeloid leukaemia, pulmonary fibrosis, cirrhosis, premature aging, skin pigmentation and nail abnormalities^6-9^.

### The role of molecular diagnostics in TBD

A genomic diagnosis for patients presenting with TBD is vital for optimal treatment. The majority of patients with TBD and bone marrow failure or a myeloid neoplasm will need an allogenic bone marrow transplant (BMT) for treatment. Currently, a matched-related donor BMT remains the treatment of choice. In the context of TBD, it is vital to exclude the presence of the causative gene variant in the donor. Patients with an underlying TBD may also present non-specifically, such as with adult-onset idiopathic pulmonary fibrosis. Detection of a causative variant identifies patients likely to respond to TBD-specific treatments^10,11^. In practice, however, less than half of suspected cases receive a genomic diagnosis. The rarity, partial penetrance, anticipation and variant spectrum (including missense, loss-of-function and non-coding variants, in addition to high gene and locus heterogeneity) of TBD limits the evidence available for variant curation^9,12,13^. Furthermore, as the majority of TBD is associated with an autosomal dominant mode of inheritance, a large number of variants of uncertain significance (VOUS) are reported to clinicians. This results in significant uncertainty about the safety of a matched-related BMT. To address this issue, there is a clinical need to reduce the number of variants requiring a VOUS classification.

### Functional genomics in TBD

Improving the ability to curate using functional genomics results (the American College of Medical Genetics (ACMG) and Association for Molecular Pathology (AMP)^13^ criterion PS3/BS3 may reduce the number of VOUS reports. As there are no diagnostic assays available for TBD genes, curators must look to the published scientific literature. The PS3/BS3 criteria guide the weighting of published functional studies performed on a given variant of interest. Published functional assays, however, rarely meet these assay validation requirements^14^. There is a need to optimise the functional assays being currently performed in research laboratories so that they address ACMG/AMP requirements for diagnostic variant curation. This review will focus on the current landscape of functional assessments of variants associated with TBD and explore assays that may meet PS3/BS3 criteria.

## Methods

### Literature review

The genes of interest were chosen based on established gene-disease validity for TBD. Both ClinGen and Panelapp data were reviewed and only genes with established gene-disease validity for TBD were included^15,16^. Published assay results for variants in the established genes were identified using the Mastermind and OpenTargets databases^17,18^ (Figure 1). Both databases use artificial intelligence and logical searches to correct for variability in gene and variant nomenclature usage^17,18^. Screening using Mastermind was performed by two independent reviewers using the imported abstracts and inbuilt highlighting of article text. If the abstract or highlighted text discussed functional studies the article was stored for retrieval. OpenTargets sorts articles based on supplied evidence for gene involvement in a biological process of interest. For each gene of interest all articles tagged with an experimental evidence code were stored for retrieval. Articles were excluded if no single nucleotide variations (SNVs) or insertion/deletions (indels) were assessed for the following genes: *ACD, CTC1, DKC1, NAF1, NHP2, NOP10, RTEL1, TERC, TERT, TINF2, STN1, WRAP53*. Nomenclature for all variants was confirmed and reported using the canonical transcripts in VarSome^19^ (supplemental Table S1). Given that few articles were identified for *PARN* and *ZCCHC8*, all functional genomic data interrogating the role of these genes in telomere biology were included. Articles reporting on animal model systems were also considered following screening for TBD related phenotypes. Correlation between the total citation count and number of published functional assessments per variant for *TERT* (Mastermind search) was determined to assess risk of reporting bias. Where possible, unpublished data from functional genomics laboratories were also included (Figure 1; manuscripts in preparation).

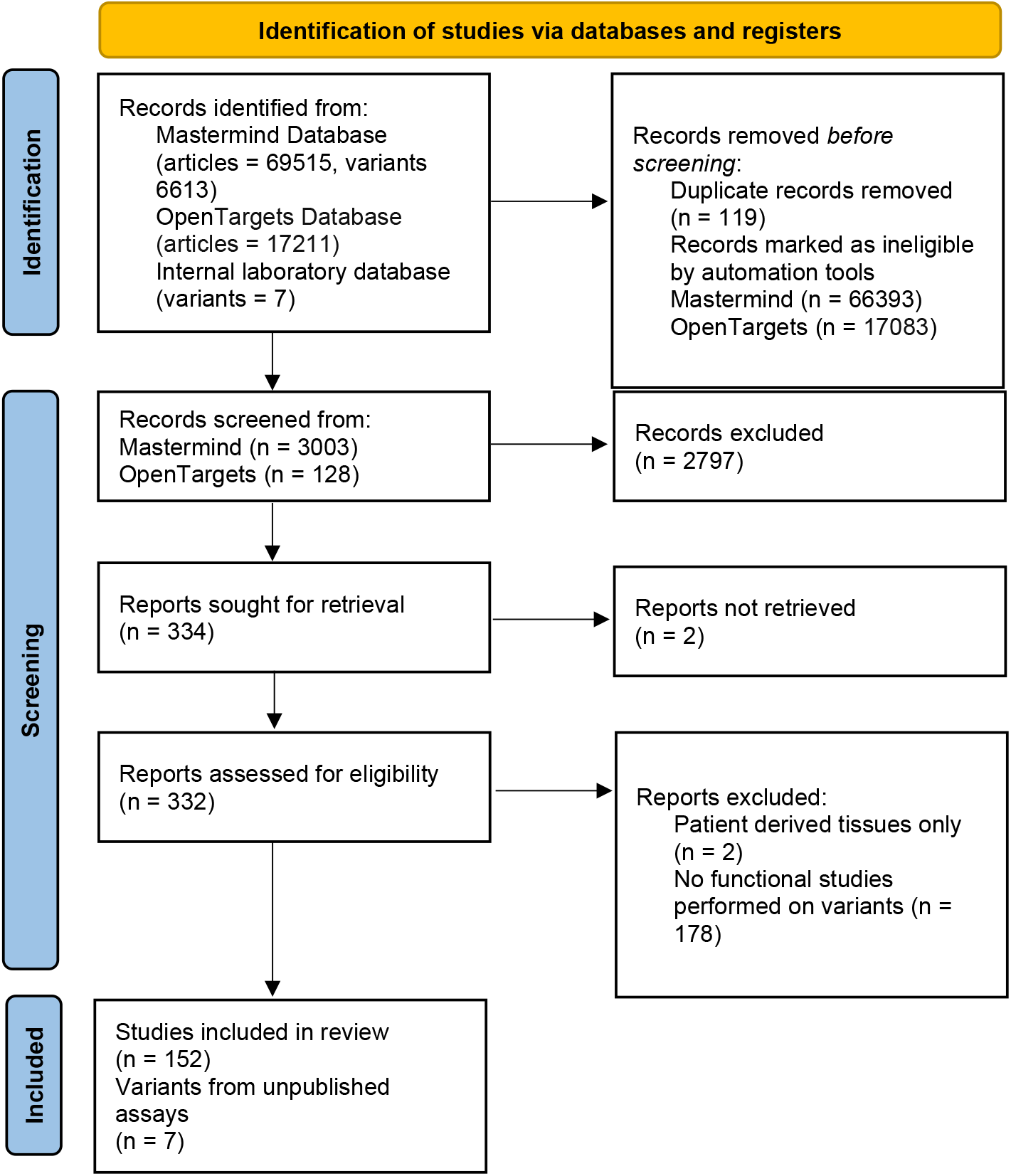

### Sensitivity and specificity calculations

In order to apply PS3/BS3 the sensitivity, specificity, positive predictive value (PPV) and negative predictive value (NPV) of an assay must be assessed. These calculations are as follows:

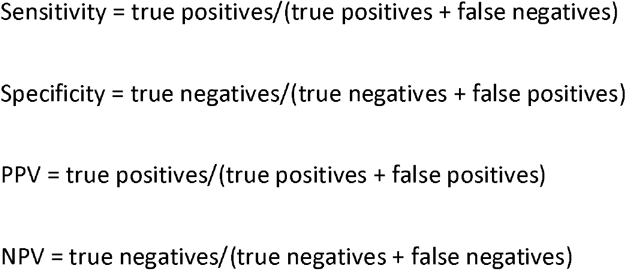

Only studies reporting on *TERT* variants yielded sufficient data to perform an assessment of sensitivity and specificity. *TERT* variants were tabulated in order from 5’ to 3’ (supplemental Table S2). Sensitivity and specificity estimate calculations were performed using *TERT* variants considered to meet ACMG/AMP criteria for likely pathogenic/pathogenic (LP/P) or likely benign/benign (LB/B).

*TERT* variants were included in the analysis if they met the following inclusion criteria

1. The variant must have been identified in a human subject (reported in VarSome OR ClinVar OR UniProt OR the published literature)^19-21^.
2. The variant was curated by the authors as LP/P or LB/B based on available data in VarSome, genome aggregation database (gnomAD) and the published literature without the application of PS3 or BS3^19,22,23^. In brief, application of ACMG curation guidelines are in line with previously published curation strategies^9^. Further details regarding curations are available in the supplemental methods.

As the cut offs for application of some ACMG/AMP criteria could be considered discretionary, a sensitivity analysis using altered criteria was performed (supplemental methods). Due to the limited amount of data available for a given functional assay, data were pooled across studies. Where results were conflicting, the underlying methodology was considered. Specifically pertaining to the DTA assay, instances using human cell lysates were considered superior to results from assays using non-human cell lysates. For qualitative assays, a non-wildtype result was considered ‘functionally abnormal’. Conversely, a result similar to the wildtype protein was considered ‘functionally normal’.

For quantitative assays such as the direct telomerase assay (DTA), a functionally abnormal result was considered as an activity level <75% of wildtype OR significantly reduced repeat addition processivity (see supplemental Figure S3).

### Odds of pathogenicity

To assess the sufficiency of numbers of controls used for the DTA and Telomerase Repeated Amplification Protocol (TRAP) assays, odds of pathogenicity (OddsPath) calculations were performed as described in the supplemental methods and elsewhere^14,23,24^. Briefly, the OddsPath is calculated using a Bayesian classifier^24^. The prior probability is determined using the number of established variants tested. The posterior probability is determined using the assay results. OddsPath ranges for evidence strength (strong, moderate and supporting) were used as recommended by ClinGen guidelines^14^.

### Application of PS3/BS3 to VOUS TERT variants

PS3/BS3 application to *TERT* VOUS variants was as follows:

PS3_supporting = Functionally abnormal DTA result.

PS3_moderate = Functionally abnormal DTA result.

AND

Functionally abnormal telomere extension assay OR fibroblast immortalisation assay.

BS3_supporting = Functionally normal DTA result

## Results

Of the 332 articles considered for eligibility, 152 articles met inclusion criteria (Figure 1). In total, functional genomics results for 407 variants from the 14 genes included in the review were identified. ACMG classifications for all variants are shown in figure 2 and reported in the supplementary data (Supplemental Tables S2, S4, S5 and S6). *TERT* was the only gene where sufficient data was available to assess assay sensitivity, specificity, PPV and NPV to enable PS3/BS3 recommendations. Data related to all other genes is presented descriptively. All published functional studies focused on testing variants with a high pre-test probability for causality. No single study published data on sufficient numbers of benign and pathogenic variants (at least 11 in total) to meet ACMG/AMP recommendations for PS3/BS3 application^14^. There was significant heterogeneity in assay choice and assay design which may have introduced bias for the pooled data. This also resulted in conflicting results for some variants. Published functional assessments did appear to correlate with the number of citations overall for a given variant, suggestive that publication bias would not have significantly impacted results (Supplemental Table S7). Overwhelmingly, studies were conducted in cells lines or using purified protein/RNA; however, 18 studies functionally validated findings in animal models (Supplemental Table S8).

**Figure 2.**
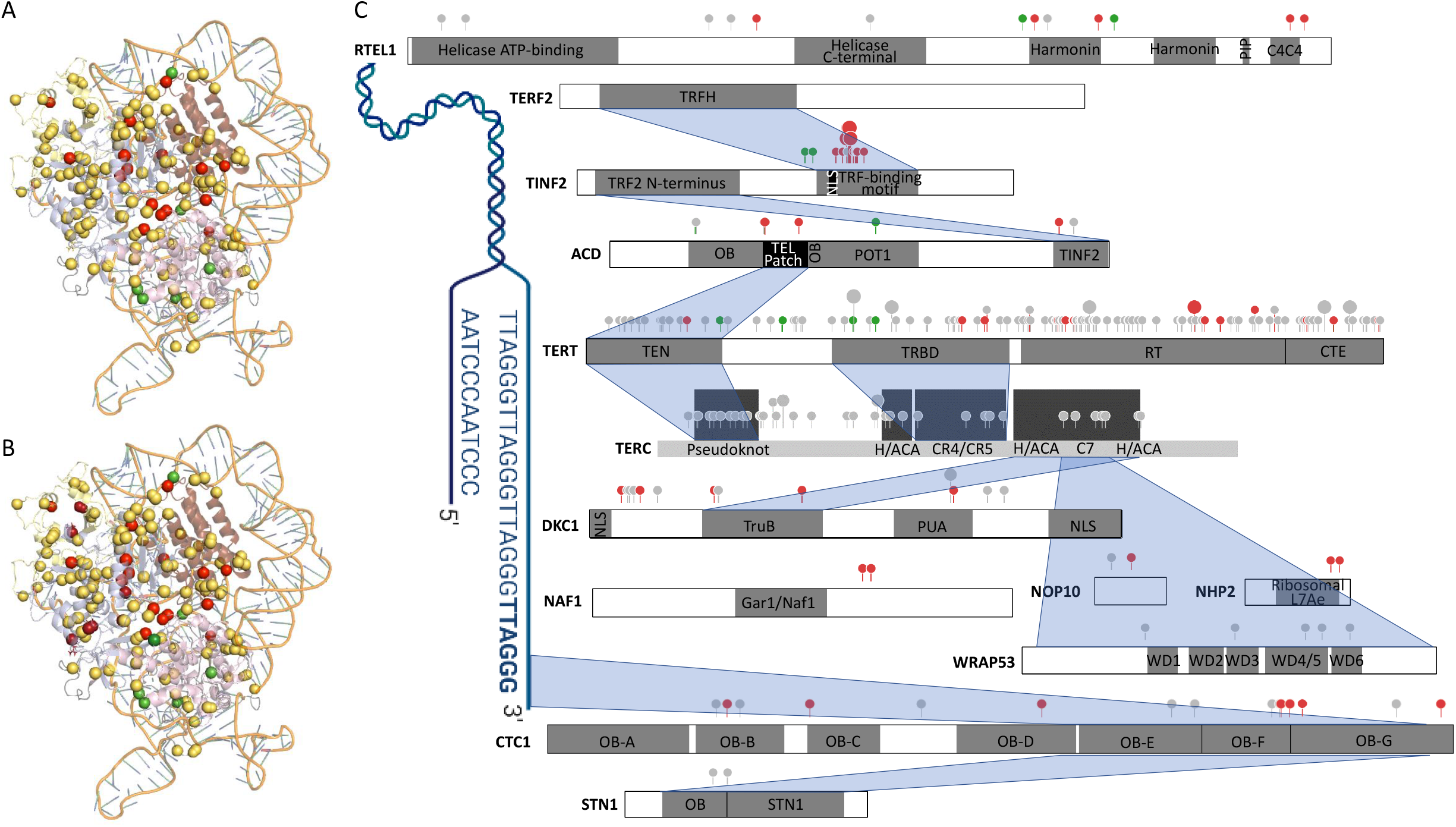
A: Location of pathogenic, VOUS and benign variants within the TERT structural model. TEN domain (pale yellow); TRBD domain (light pink); RT domain (light blue); CTE (chocolate); Linker (light grey); Pathogenic and likely pathogenic variants (red sphere); benign and likely benign variants (green sphere); VOUS (light orange sphere); hTR RNA (orange strand). Adapted from The PyMOL Molecular Graphics System, Version 1.2r3pre, Schrödinger, LLC. B: Location of pathogenic, VOUS and benign variants within the TERT structural model following reclassification using proposed PS3/BS3 rules. Variants reclassified to pathogenic/likely pathogenic shown in dark red, variants reclassified to benign/likely benign shown in forest green. Adapted from The PyMOL Molecular Graphics System, Version 1.2r3pre, Schrödinger, LLC. C: Summary of TBD gene variants with published functional genomic data. PIP (PCNA interaction motif), TRFH (telomeric repeat-binding factor homology domain), TRF (telomeric repeat-binding factor), NLS (nuclear localisation signal), TRF2 (telomeric repeat-binding factor 2), OB (Oligonucleotide/Oligosaccharide-Binding fold), TEN (essential N-terminal domain), TRBD (the RNA binding domain), RT (reverse transcriptase domain) (RT), CTE (C-terminal extension), H/ACA (H/ACA box), PUA (PseudoUridine synthase and Archaeosine transglycosylase domain), WD (WD-repeat domain). Pathogenic and likely pathogenic variants represented in red, variants of uncertain significance are represented in grey and likely benign and benign variants are represented in green. Estimated protein domain interactions represented in blue. Adapted from “Telomere repeats”, by BioRender.com (2022). Retrieved from https://app.biorender.com/biorender-templates and ProteinPaint^133^.

### TERT

*TERT* encodes for the TERT protein, the catalytic subunit of telomerase. Telomerase is the enzyme complex responsible for addition of DNA to telomeres^25^. The telomerase complex also consists of the proteins dyskerin, NOP10, NHP2, GAR1, TCAB1 and the template RNA hTR^1-3^. Unlike the other vital components of the telomere maintenance system, *TERT* expression is limited to stem cell populations^26-28^. *In vitro*, transfection of the *TERT* gene is sufficient to prevent the normal senescence of mortal cells such as fibroblasts^29^. We identified 137 *TERT* variants in patients with a wide range of phenotypic presentations including Hoyeraal-Hreidarsson syndrome, dyskeratosis congenita, idiopathic pulmonary fibrosis, cirrhosis and myeloid malignancy that have been assessed using functional assays (supplemental Table S2). Of those, 23 variants had sufficient information published to enable a non-VOUS classification using ACMG/AMP guidelines (table 1, supplemental Table S9). The assays used for functional testing could be grouped into four broad categories:

1. Telomerase enzymatic activity of purified telomerase or cellular lysates (using either the non-PCR-based DTA and or the PCR-based Telomere Repeat Amplification Protocol (TRAP))^30,31^.
2. Telomere elongation potential when introduced into cultured cells (Telomere elongation assay (TEA)).
3. Protein-nucleic acid interaction studies such as telomere localisation.
4. Fibroblast immortalisation potential (measured using endpoints such as mean population doublings achieved, or bypass of senescence (measured by detection of senescence-associated β-galactosidase activity or Telomere Dysfunction Induced Foci analysis))^32,33^.

**Table 1.**
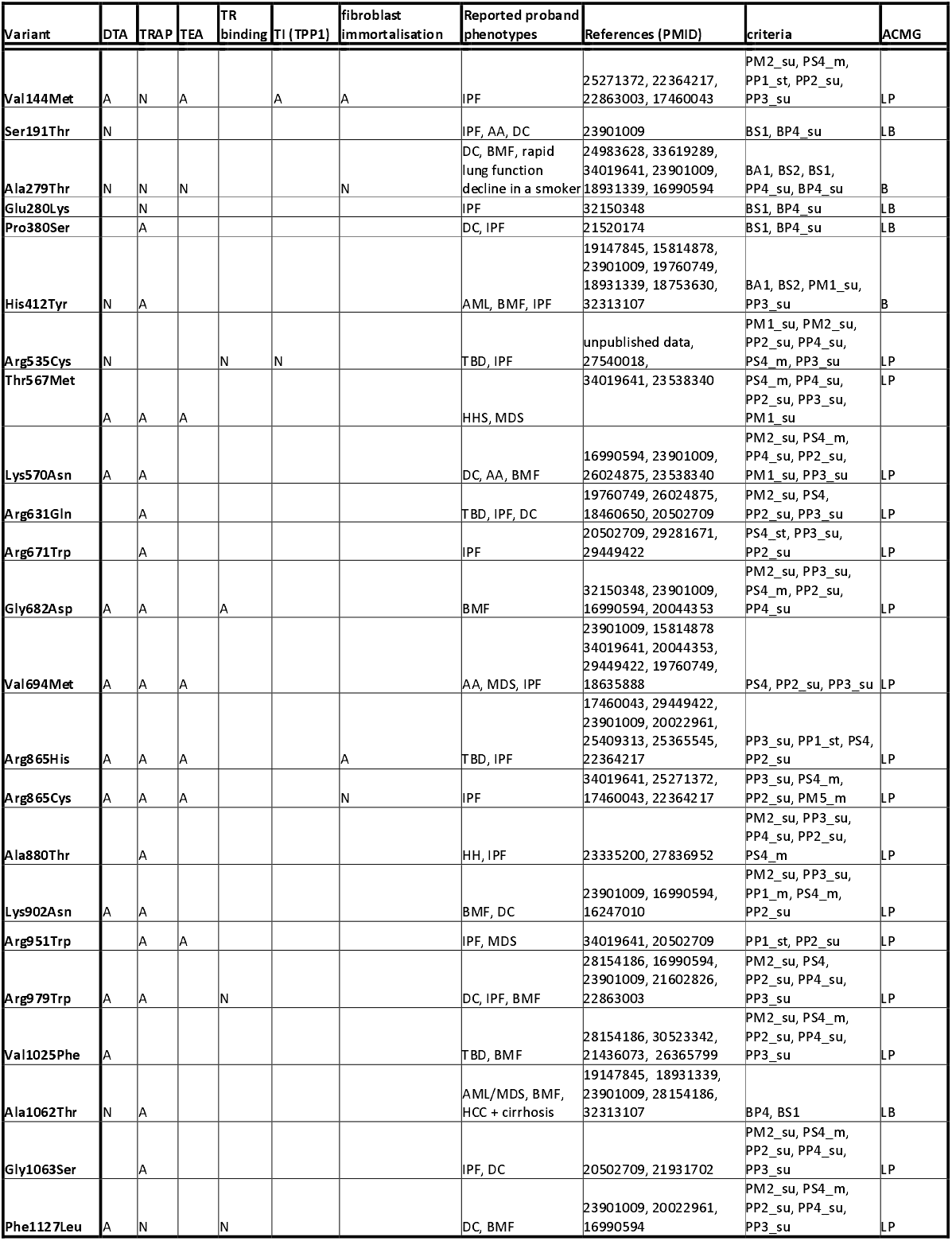
Non-VOUS TERT variants with functional genomic assay results.

The most extensively used assays were the DTA and TRAP. The DTA was estimated to have a sensitivity of 92% and specificity of 100%. The OddsPath for PS3 and BS3 for the DTA was 3.7 (supporting strength) and 0.067 (moderate strength) respectively. TRAP was associated with a sensitivity of 87% and specificity of 40% and OddsPath of moderate strength for BS3 (supplemental Tables S10, S11). OddsPath score for PS3 application was not reached for the TRAP assay (supplemental Table S11). Insufficient data were available to assess the other assays. A sensitivity analysis using alternative PM1 and PS4 criteria for TERT variant classification was also performed (supplemental Table S12). Modest changes were seen; however, the sensitivity for the DTA remained high and the specificity for the TRAP assay remained low.

Along with telomerase activity, a measure of the ability of telomerase to lengthen telomeres, such as the TEA or fibroblast immortalisation, may also be required as the full spectrum of telomerase functional requirements are not captured by activity assays alone. For example, the Val144Met variant was initially associated with wildtype telomerase activity using DTA and TRAP^34^. This variant is suspicious for causation as it is found in the TEN domain, absent from gnomAD, curated 4 times in ClinVar and more than 7 segregations in affected family members reported in two separate families^19,21,22,35^. In keeping with the clinical findings, impaired fibroblast immortalisation has been reported^34^. Following the initial studies, it was found that this discrepancy is due to Val144Met causing a POT1-TPP1 binding defect and reduced telomerase recruitment to telomeres^36^. This highlights the limitations of isolated protein assays. TEA or fibroblast immortalisation assays may also assist in accounting for unidentified regions of TERT that are important for telomerase regulation. Based on the above findings, PS3/BS3 was applied to the 114 VOUS *TERT* variants and 9 were reclassified (8 to LP and 1 to LB; supplemental Table S13).

### TERC

The telomerase RNA component (hTR), encoded by *TERC*, is a 451 nucleotide lncRNA that provides the template for telomere extension by telomerase^37,38^. Adequate levels of hTR are limiting for telomerase activity^39^ and hTR’s interaction with TERT and telomeres is dependent on a complex tertiary structure of hair-pin loops like other box H/ACA small nucleolar RNAs. Important structural domains include the template/pseudoknot and CR4/CR5 domains (mediating TERT interaction) and the H/ACA domain (essential for hTR accumulation)^40-43^. The H/ACA ribonucleoprotein (RNP) complex (dyskerin, NOP10, GAR1, NHP2) interacts with the H/ACA domain of hTR and regulates rate of hTR degredation^37,43^.

ACMG/AMP rules are challenging to apply to *TERC* variants, as *TERC* does not encode for a protein product. To evaluate functional assays, an alternative PS4 criterion was used to identify variants most likely to be pathogenic (supplemental methods). In an analysis combining both *TERT* and *TERC* variants, the DTA appeared potentially useful for *TERC* variant assessment (supplemental Table S14). As with *TERT* variants, it is likely that correlation with orthogonal methods such as telomere elongation potential will also be useful, particularly for variants that impact hTR localisation^44^. Other auxiliary assays used for functional evaluation include co-immunoprecipitation to measure binding to TERT and hTR transcript quantification (supplemental Table S4)^45-48^. These assays may play a role in the functional assessment of variants involving relevant functional domains.

### ACD

*ACD* encodes for the protein TPP1, a member of the shelterin complex. The shelterin complex (TPP1, TIN2, TRF1, TRF2, POT1 and RAP1) caps the ends of telomeres to prevent recognition as double strand breaks^49^. Dysfunction of the shelterin complex can result in triggering of cell senescence, non-homologous end joining, interchromosomal fusions and unregulated telomerase activity depending on the functionality of cell cycle checkpoints^50-52^. This protective telomere cap is dependent on stabilisation of TIN2 on telomeres by interaction with TPP1 through the TIN2 binding domain^53^.

Systematic site-directed mutagenesis has shown that variants disrupting TIN2 binding are associated with increased DNA damage marker localisation to telomeric DNA and hematopoietic stem and progenitor cells (HSPCs) engraftment failure in irradiated mouse models^54^. While these assays may be useful in evaluating TIN2-binding domain variants, only a single variant, the Pro491Thr (reported *in trans* with Lys170del in a case of Hoyeraal-Hreidarsson syndrome) has been reported in this domain^55^.

The Oligonucleotide/Oligosaccharide-Binding (OB) fold of TPP1 also promotes telomere elongation by encouraging TERT recruitment to telomeres through a direct interaction between the ‘TEL patch’ of TPP1 and the TEN domain of TERT as well as by enhancing telomerase processivity^56-60^. Given that the presence of recombinant TPP1 is known to indirectly enhance telomerase processivity, TRAP and the DTA have been used to assess multiple *ACD* variants in the TEL patch and OB fold. The Pro507Leu, Glu169del and Lys170del variants have shown reduced binding to telomerase and/or telomerase processivity enhancement^55,61-63^. Auxiliary assays assessing telomerase recruitment to telomeres such as fluorescence *in situ* hybridisation probe localisation of hTR and telomeres may also aid in OB fold and TEL patch variant assessment^55,64^. mRNA expression levels are not likely to be helpful in assessing *ACD* variants as protein upregulation appears to compensate for copy number loss or transcripts that undergo nonsense mediated decay^63^.

### CTC1 + STN1

The CTC1 protein forms a component of the Cdc13/Ctc1, Stn1, Ten (CST) complex, together with STN1 and TEN1. By recruitment of DNA pol α-primase, the CST complex facilitates fill-in DNA synthesis of the lagging strand (the C-strand) to prevent telomeric shortening^65-69^. Of the CST complex genes, *CTC1* has been the most extensively studied (supplemental Table S5). Patients presenting with TBD show compound heterozygosity for a loss-of-function variant and a hypomorphic variant^70^. While pathogenic *CTC1* variants are associated with varied mechanisms of pathogenesis, the outcome is impaired C-strand fill-in^71-73^ (supplemental Table S5). Detection of C-strand fill-in failure is likely to be the most useful functional assay for functional assessment of *CTC1* and possibly *STN1* variants. Quantification of single stranded telomeric DNA (a G-overhang) using oligonucleotide probe hybridisation in native gels has demonstrated promising discrimination of pathogenic *CTC1* variants from a wild-type control^73^ (supplemental Table S5). The notable exception was the preserved fill-in associated with the Leu1142His variant. This was likely due to masking by presence of wildtype *CTC1* protein, highlighting that assays designed with heterozygous cell lines may lead to false negative results^74^. The functional assessment of *STN1* variants is limited. Patient-derived fibroblasts harbouring the variants Arg135Thr and Asp157Tyr both showed reduced population doubling and increased senescence and chromosomal fusions compared with healthy controls^75^. This was associated with increased G-overhang as well as non-telomeric effects (reduced recovery from hydroxyurea-induced replication fork stalling)^75^.

### DKC1 + NAF1 + NOP10 + NHP2

Dyskerin, NOP10 and NHP2 are the non-catalytic components of the telomerase ribonucleoprotein complex and are important for maintaining adequate hTR levels for telomere extension^43,76,77^. NAF1 is also a member of the hTR binding ribonucleoprotein complex but it is replaced by GAR1 once the complex is localised to the Cajal body^78^. Of the proteins in the ribonucleoprotein complex, *DKC1* has been the most extensively studied. Putative pathogenic variants have been associated with impaired ribonucleoprotein complex assembly (Thr66Ala and Met350Thr) or more commonly, impaired hTR binding (Gln31*, Leu37del, Lys39Glu, Lys43Glu, Thr49Met, Ser121Gly, Met350Ile, Ala353Val, Gly402Glu)^45,79-81^, resulting in increased hTR polyadenylation and increased hTR degradation^80,82^, which is supported by studies from patient-derived cells that have shown reduced hTR levels compared to wild-type controls^43,83,84^. Functional assessment of hTR levels would be a rapid and relativity simple functional assay if shown to have acceptable sensitivity and specificity with further studies of *DKC1* variants in mutant cell lines.

Multiple other functional assays have also been used to assess *DKC1* variants. The Glu206Lys and Ala353Val variants show reduced telomerase activity measured by TRAP ^85^. The Glu206Lys variant has been associated with a similar haematopoietic defect phenotype to a *DKC1* null zebrafish and impaired phenotypic rescue following mRNA injection^86,87^. While exon 15 deletion (corresponding to a 2kb deletion reported in a family expressing a TBD phenotype) is associated with increased DNA damage sites and accelerated HSPC loss in mice, those carrying the Ala353Val or Gly40Glu variants have not shown a TBD-like phenotype^88-90^.

In addition to their roles in telomerase activity, Dyskerin and NOP10 also have a role in pseudouridylation of ribosomal RNA^87,91,92^. Reduced ribosomal RNA pseudouridylation has been reported in patient-derived cells harbouring *DKC1* variants^91,92^. Variants thought to preferentially impact the pseudouridylation catalytic site (*DKC1*:Glu206Lys and *NOP10*:Thr16Met) may be associated with non-TBD phenotypic features such as nephrotic syndrome, cataracts and sensorineural deafness^87^. These findings are suggestive that functional assay selection may depend on patient phenotypic features and variant location within the protein.

Knockdown of NOP10 reduces hTR levels by approximately 90%^93^ Rescue with a *NOP10* Arg34Trp-expressing plasmid did not improve hTR levels in comparison to a partial restoration conferred by wildtype NOP10 transfection^93^. Unlike the majority of *DKC1* variants, there is some evidence that variants in *NOP10* may interfere with H/ACA RNP-complex assembly. Purified recombinant protein with the Arg34Trp variant was shown to impair pre-ribonucleoprotein assembly^45,94,95^. This pre-ribonucleoprotein assembly assay was also performed for the *NHP2* variants Val126Met and Tyr139His and showed impaired pre-ribonucleoprotein assembly as well as impaired NOP10 binding^94,95^. Functional data for *NAF1* is limited to hTR expression data. Reduced hTR expression in HCT116 cells has been shown for the Lys319fs*21 and Ser329fs*12 variants compared to wildtype^94^.

### PARN

*PARN* encodes poly(A)-specific ribonuclease. It is required for hTR biogenesis. Loss of PARN function results in approximately 30% of the normal mature level of hTR in patient-derived fibroblasts from individuals with TBD and an associated increase in transcripts with 3’ oligo(A) tails which target hTR for degradation^96-98^. Acetylation of the Lys-566 residue is known to alter PARN activity and also adenylated hTR levels^99^. Given the rarity of *PARN* variants, a functional assay using known LB/B and LP/P variants would be challenging to validate. Quantification of hTR levels, if further validated in other genes such as *DKC1*, may provide functional evidence for *PARN* variants.

### RTEL1

The RTEL1 protein is a DNA helicase which assists in the resolution of secondary structures of DNA, thus modifying DNA-protein interactions^100^. Given this broad role, the function(s) most relevant to TBD has been difficult to clarify. Inducible expression of wildtype RTEL1 in patient-derived cell lines was suggestive that maintenance of the G-overhang may be the most important function^101^. RTEL1 (along with TRF2) is thought to assist the G-overhang to tuck into a protective loop of double-stranded telomeric DNA (the t-loop)^102^. G-overhang loss has been associated with six out of the ten TBD patient-derived samples assessed (Val1294Phe, Met492Ile/Arg974*, Pro82Leu, Met652Thr, Pro867Leu and Pro884Leu)^101,103-105^. Of these, Pro867Leu is a LB variant (allele frequency 0.0007483 in European (non-Finnish gnomAD v2.1.1^106^ and REVEL score 0.009999^107^)^105^. Of the four samples showing relatively preserved G-overhangs, one sample harboured Pro996His, a LB variant^103^. G-overhang loss can be quantified using in-gel hybridisation a similar approach to c-strand fill-in assessment for *CTC1* variants^101^. Assays interrogating RTEL1-dependent resolution of DNA secondary structures have also revealed alterations associated with an *RTEL1* variant (Lys48Arg) and may also be useful^108^.

Markers of chromosomal damage such as γ H2AX staining have also been associated with RTEL1 loss-of-function; however, in studies of patient-derived cell lines (Trp456Cys, Ile425Thr,

Cys1244Profs*17, Pro884_Gln885ins53*13, and Val796Alafs*4, Arg974* and Met492Ile, Arg1264His, Cys1279Ala/Cys1282Ala) results were varied^105,109-111^. Given the broad functions of RTEL1, these phenotypes may not necessarily be related to defective telomere maintenance, which may limit their application. Domain-specific functional assays may be useful as ancillary tests for certain variants. C4C4 domain variants were associated with reduced TRF2 binding in a pull-down assay^110^.

### TINF2

TIN2 is a shelterin complex protein that has positive and negative impacts on telomere elongation. It enhances processivity by recruiting telomerase and POT1/TPP1 to telomeres yet also recruits TRF1 to telomeres which inhibits telomere elongation^112-114^. These multiple roles may explain why demonstration of functional consequences of *TINF2* variants has been difficult even though all causative variants are found in a discrete region of the gene (the DC cluster)^115^. Studies assessing TIFs, protein localisation and hTR expression have not demonstrated functional defects associated with DC variants^56,60,116^. Analyses of binding between TIN2 and its protein partners have had mixed results and suggest co-immunoprecipitation studies are likely insufficient for PS3/BS3 application. While the Gln269* variant disrupts TIN2-TRF1 interaction, Arg282His (the most commonly reported causative variant in *TINF2*) does not impact protein interaction^117,118^. A published assessment of 20 putative causative *TINF2* variants also did not show changes in TRF1, TRF2 or TPP1 binding^119^. These findings may be due to differences in TIN2 isoforms. There are three known isoforms of TIN2 (TIN2S, TIN2L and TIN2M), all of which contain the DC cluster and have been shown to localise to telomeres but may have differing interactions with TRF1 and TRF2^56,120^. Co-immunoprecipitation studies have shown that the Arg282His reduces TIN2L-TRF2 interaction but not TIN2S-TRF2 interaction^120^. In the HT1080 cell line, TIN2S overexpression resulted in telomere lengthening inhibition, whereas TIN2L overexpression did not show this effect. The Arg282His variant of either isoform suppressed telomere lengthening, suggesting that DC cluster variants may result in TIN2L acquiring a TIN2S-like inhibitory effect on telomere lengthening.

As TIN2 enhances telomerase processivity and recruitment to telomeres via TPP1/POT1^56,60,116^, telomerase activity studies and TRF analysis have also been used to investigate the functional effect of *TINF2* variants. TRF analysis demonstrated an increased rate of telomere attrition associated with the Lys280Glu, Arg282Ser and Arg282His compared to wildtype with all three missense variants^121^. The TRAP assay however, did not reproducibly show a defect in telomerase activity for these variants. Given that TIN2 is associated with increased telomerase processivity, the DTA may be more sensitive^56,121^. Alternatively, telomerase activity can be measured after TIN2 pulldown, as a measure of TIN2 interaction with telomerase; all three of the above-mentioned variants showed defects in telomerase association.

Other DC cluster variants have been associated with defective telomere cohesion during S phase which also may impact telomere length^122^. Residues 283 to 287 (PTVML) of TIN2 correspond to a binding site for HP1γ, that is required for sister telomere cohesion^122^. Binding of *TINF2* to HP1γ was reduced and telomere cohesion altered for the Arg282His, Lys280*, Pro283His, and Leu287Pro compared with wildtype, however, not all DC cluster variants reduced HP1γ binding^122^.

### WRAP53

TCAB1, encoded by *WRAP53*, interacts with hTR and other RNAs of the scaRNA family^123,124^. Depletion or knock-out of TCAB1 has implicated it in localisation of telomerase to Cajal bodies and telomeres^46,47,123^, correct folding of hTR^47^, and telomere maintenance^47,123,125^. Transfection of the Phe164Leu, Leu283Phe, Arg398Trp, His376Tyr and Gly435Arg variants resulted in abnormal localisation of hTR to the nucleolus as well as reduced levels and abnormal folding of the TCAB1 protein^126-128^. Similarly, studies of patient-derived cells and knock-out HeLa cell lines also showed reduced localisation of hTR to the Cajal bodies^47^. Given the recurrently reported hTR localisation defect, hTR localisation studies may provide a useful functional assessment of *WRAP53* variants.

TCAB1 also has non-telomeric roles, which has been proposed as a possible explanation for the severe neurological deficits associated with *WRAP53* variants^127^. Assessment of non-telomeric DNA protective functions of TCAB1 may therefore be useful. Detection of DNA damage sites may also provide functional details about *WRAP53* variants as TCAB1 also accumulates at DSBs with H2AX and facilitates RNF8 accumulation^129^.

### ZCCHC8

*ZCCHC8* is a member of the nuclear exosome-targeting (NEXT) complex^130^. The NEXT complex is important for degradation of non-coding RNA. There is little published functional genomics data available for *ZCCHC8*. The sole variant so far identified in TBD patients, Pro186Leu, has been associated with a 50% reduction in ZCCHC8 protein compared with wildtype in primary fibroblasts or when introduced into human cell lines^5^. ZCCHC8 depletion in human cells has also been associated with reduced levels of hTR^5^. This may mean that hTR quantitation may also assist with *ZCCHC8* variants. A heterozygous knock-in mouse model also showed reduced TR levels; however, a haematological defect was not seen^5^. Null mice demonstrated a severe neurodevelopmental phenotype^5^.

## Discussion

Development of an evidence-based guideline for applying PS3/BS3 to variant curations for TBD genes may resolve a substantial number of VOUS variants. The data summarised in this review suggest that the DTA is currently the assay of choice for assessing *TERT* variants. As the results presented in this review were collated from non-standardised assays, the strength level was limited to supporting level only^14^. Encouragingly, the OddsPath score for the DTA was within the range and close to for BS3_moderate and PS3_moderate strength respectively, suggestive that further results from a single standardised DTA would allow PS3 and BS3 to be applied at a moderate strength level. Applicability to other genes such as *TERC, DKC1, NAF1, NOP10* and *NHP2* may also be achievable with a single standardised DTA. An assay assessing the phenotypic impact of reduced telomerase activity such as the TEA or fibroblast immortalisation studies may resolve conflicting findings and may also allow for a more granular grading of PS3/BS3 (supporting, moderate and strong levels).

Results from this study are suggestive that the TRAP assay has poor specificity (40%) and therefore we do not recommend its use for PS3. The TRAP assay is non-quantitative and does not assess processivity; these limitations may explain these findings^131,132^. The DTA appears more sensitive then the TRAP (92% compared with 87%) and thus the DTA is preferred for applying BS3. Based on the current data the following recommendation for PS3/BP3 application for *TERT* variants is proposed.

PS3_supporting: DTA activity < 75% of wildtype control.

PS3_moderate: DTA activity < 75% of wildtype control.

AND

TEA reduced OR fibroblast immortalisation impaired.

BS3_supporting: DTA activity > 75% of wildtype control.

### Limitations

Of the 14 genes reviewed, only 10 LB/B variants (including 6 *TERT* variants) have published functional assessments. This limited the specificity and NPV assessments to *TERT* only. Of the LB/B variants assessed, they all were included in functional assays due to a suspicion for causality. No published literature specifically assessing established benign variants was found. As such, the minimum level of protein function required for normal cellular function remains unconfirmed for all TBD related genes. A second significant limitation is the lack of standardisation of the commonly used assays such as the DTA. This results in reduced reproducibility and may explain the conflicting results for some variants. Literature searches for gene variants is challenging due to alternative variant and gene nomenclature and differences in reference transcript choice. While the data mining algorithms used in Mastermind and OpenTarget attempt to account for this, it is possible that published functional studies may have been missed due to differences in gene and/or variant nomenclature.

### Improving the strength of evidence for PS3/BS3

The understanding of the biology of TBD is increasing rapidly and functional genomics research has been a significant driver of this. As the vast majority of historical studies of variant function have not been designed for the purposes of PS3/BS3 classification there are a number of recurring limitations which often preclude PS3/BS3 application. These include a lack of inclusion of established benign variants, lack of assay standardisation and limited replication studies. To improve the applicability of functional genomic data for PS3/BS3 application the following assay design considerations were identified:

1. Standardisation of the DTA and replication testing of previously assessed established LP/P and LB/B variants.
2. Evaluation of additional established LB/B variants.
3. Evaluation of the utility of a dual assessment using DTA and telomere elongation.
4. Consideration of the DTA for assessment of *TERC, DKC1, NAF1, NOP10, NHP2* and *PARN* variants.
5. hTR quantification may be a fast and simple assay to assess *TERC, DKC1, NAF1, NOP10, NHP2 PARN* and *ZCCHC8* variants.
6. In-gel hybridisation interrogation of the G-overhang may assist with classification of *CTC1, STN1* and *RTEL1* variants.
7. Telomere recruitment assessment using hTR/telomere FISH may be useful for *ACD, TINF2* and *WRAP53* variants.
8. Selection of a subset of variants and introduction into an established animal model (e.g. zebrafish for *STN1* and RNP complex associated genes) would further correlative evidence for variant classification.

## Conclusions

The body of functional genomic research performed to understand the molecular biology of TBD is substantial and has been developed over many decades. Unfortunately the use of PS3/BS3 is currently limited by assay heterogeneity, lack of correlation studies and paucity of LB/B variant assessment. There appears, however, to be significant potential for the established assays to be incorporated into a workflow that would strengthen the PS3/BS3 criteria. Given the complexity of telomere biology, it is likely that assessment would require a combination of telomerase activity and telomere lengthening potential. To achieve this, it is essential that diagnostic and research laboratories collaborate to ensure assays being developed for other indications, such as gene and variant discovery, drug target discovery and therapeutic response can also be utilised for ACMG classification.

## Supporting information

Supplemental results

PRISMA abstract check list

PRISMA check list

## Data Availability

Additional data supporting the findings of this study are available in either the manuscript and/or supplemental material.

## Acknowledgments

N.N. is supported by an Annie Bishop doctoral degree scholarship in cancer research. S.F. was funded by the Olympia Morata Program of the Medical Faculty Heidelberg. J. K. H. is funded by an RMIT VC Fellowship. K.A.F. is supported by the Alex Gadomski Fellowship, funded by Maddie Riewoldt’s Vision. J.D. is supported by a Select Foundation Fellowship. T.B. was supported by the Arcus Foundation and the Hill Foundation.

## Author Information

Conceptualization: N.N., K.F.; Data curation: N.N., A.A., T.B., S.F.; Formal analysis: N.N., A.A., T.B., S.F., J.T., K.H.; Investigation: N.N., A.A., T.B., S.F.; Methodology: N.N., S.F.; Resources: A.A., T.B. J.T, K.H.; Software: K.F., A.A., T.B., J.T., K.H.; Supervision: J.D., K.F., S.L.; Validation: N.N., S.F., T.B.; Visualization: N.N., A.A., T.B., K.F; Writing-original draft: N.N.; Writing-review & editing: N.N., S.F., T.B., K.F., S.L., J.D.

## Ethics Declaration

All genetic data analyzed and reported in this study have been previously published, are discussed in articles that are in press and/or stem from publicly available databases such as ClinVar (https://www.ncbi.nlm.nih.gov/clinvar/). No institutional review board or research ethics committee approval was required.

## Additional Information

The online version of this article contains supplemental material, which is available to authorized users.

---

Table 1. Legend

DTA (direct telomerase activity), TRAP (Telomere Repeat Amplification Protocol), TEA (telomere elongation assay), TRF (Telomere restriction fragment length), TR (telomerase RNA), TI (Telomere interaction), PMID (Pubmed ID), ACMG (American College of Medical Genetics), A (functionally abnormal), N (functionally normal), LP (likely pathogenic), LB (likely benign), DC (dyskeratosis congenita), BMF (bone marrow failure), MDS (myelodysplastic syndrome), AML (acute myeloid leukaemia), IPF (idiopathic pulmonary fibrosis), HH (Hoyeraal-Hreidarsson Syndrome), TBD (telomere biology disorder), HCC (hepatocellular carcinoma).

