## Supplemental methods for "Functional genomics for curation of variants in telomere biology disorder associated genes, a systematic review"

#### ***TERT***

#### **PATHOGENIC CRITERIA**

The following application of ACMG criteria was applied^1^.

#### **PVS1 - Null variants**

- Standard flowchart for frameshift and nonsense^2^.
- Isoforms: Only the full-length isoform spanning 16 exons retains catalytic activity^3^. Twenty-two other isoforms have been reported due to alternative splicing^3^. Normal tissues predominantly express full length TERT and TERT_238.6, an isoform that lacks exons 1, 2, 7, 8, 11 and contains an insertion of 386bp prior to exon 3^3^. The TERT_238.6 likely undergoes NMD and is not associated with a protein product^3^. Four other splice variants have been associated with protein products and expression in cancer cells (alpha, gamma, beta, alpha-beta-gamma) and while it seems likely these proteins lack catalytic activity, they may still have non-catalytic functional activity^3^. It appears they also may regulate the activity of the full length isoform by competitively binding Telomerase RNA (TR)^4^. Note exon 7 and 8 are highly constrained suggesting that evolutionary fitness is severely impacted by loss of catalytic activity even if non catalytic functions are maintained.
- 1132aa long protein.
- All exons are associated with an important functional domain.
- Penultimate exon: 15.
- Most 3’ LOF variant reported as pathogenic is the p.Cys998Alafs*50 (ClinVar RCV000627447, no disease context).
- NMD predicted for frameshift or nonsense variants occurring up to about aa1049 (50 bp upstream from the end of exon 16).
- CTE aa936-1132.
- Next Met at codon 636. 17 pathogenic/likely pathogenic variants upstream of closest potential in-frame start codon (amino acid 1113)^5^.

**PVS1 very strong:** up to aa1049.

**PVS1 strong:** aa1050-1132.

#### **PS1 - Established Pathogenic Variant**

- Apply only if the nucleotide is different but the amino acid is the same. The variant must have been seen in the germline.
- ‘Established’ is defined as a variant that has been seen and classified as LP/P in an inhouse database OR has been curated from evidence in the literature and LP/P classification has been reached.

**PS1 strong:** applied for alternative nucleotide (nt) change for established pathogenic variant.

**PS1 moderate:** applied for alternative nt change for established likely pathogenic variant.

#### **PS2 - Confirmed de novo variant**

- Also see PM6
- Apply as usual, but likely to be rarely applicable due to age of onset of phenotype. Paternity confirmation by SNP array or SNP assessment of NGS sample (needs to be specifically stated).
- See: <https://clinicalgenome.org/site/assets/files/3461/svi_proposal_for_de_novo_criteria_v1_1.pdf>

Points per de novo occurrence where paternity is confirmed:

| **Phenotypic consistency** | **Points per proband (mutually exclusive selection)** |
| --- | --- |
| Two or more phenotypic features of a TBD | 2 |
| One phenotypic feature of TBD | 0.5 |

| **PS2_Supporting** | **PS2_moderate** | **PS2_strong** | **PS2_VeryStrong** |
| --- | --- | --- | --- |
| 0.5 | 1 | 2 | 4 |

#### **PS3 - Functional studies support damaging effect** – not applicable as under study

#### **PS4 - Enrichment in cases**

Based on odds ratio calculations performed for AML/MDS^6-8^.

- 1 proband in the published literature AND <2 alleles in gnomAD: PS4 supporting.
- 2-3 probands in the literature AND <2 alleles in gnomAD: PS4 moderate.
- 4 or more probands in the literature AND <2 alleles in gnomAD: PS4 strong.

#### **PM1 - Mutational hotspot/functional domain**

- PM1 supporting can be applied to the TRBD domain^9^.

#### **PM2 - Absent from population studies**

- Supporting level only. Zero times in gnomAD^10^.

**PM3 - Detected in trans** – not generally applicable

#### **PM4 - Protein length changes**

The following applies to single aa insertions or deletions (see PM1 for more info):

- Apply PM4 moderate if in-frame variant occurs in a functional domain of *TERT.*
- PM4 not applicable if in-frame variant occurs outside of these regions.

#### **PM5 - Novel missense change**

- Novel missense change at an amino acid residue where a different missense change determined to be pathogenic has been seen before.
- The variant must have been seen in the germline. Full curation must be performed on the previously identified variant (see PS1).

**PM5_strong:** Missense change at the same residue where ≥2 different missense changes have previously been determined to be Pathogenic. PM5_strong cannot be applied together with PM1.

**PM5_moderate:** Missense change at the same residue where a different missense change has previously been determined to be Pathogenic.

**PM5_supporting:** Missense change at the same residue where a different missense change has previously been determined to be Likely Pathogenic.

PM5 cannot be applied in combination with PM1.

#### **PM6 - Assumed de novo**

- Also see PS2
- Points per de novo occurrence where paternity is unconfirmed:

| **Phenotypic consistency** | **Points per proband (mutually exclusive selection)** |
| --- | --- |
| Two or more phenotypic features of a TBD | 1 |
| One phenotypic feature of TBD | 0.25 |

| **PM6_Supporting** | **PM6_moderate** | **PM6_strong** | **PM6_VeryStrong** |
| --- | --- | --- | --- |
| 0.5 | 1 | 2 | 4 |

#### **PP1 - Co-segregation with disease**

|  | **PP1 strong** | **PP1 moderate** | **PP1 supporting** |
| --- | --- | --- | --- |
| Phenotype | One or more phenotypic features of a TBD | One or more phenotypic features of a TBD | One or more phenotypic features of a TBD |
| Number of meioses | 7 or more | 5-6 | 3-4 |

- Enumeration based on literature search (google and mastermind^11^) and inhouse database.
- Meioses within and between families weighted the same.
- Only include genotype positive and phenotype positive meioses and obligate carriers.
- Do not include the proband/s in the count.
- Cases of parent-child only literature reports: this can be considered as 1 meioses.

#### **PP2 - Missense variants are a common mechanism of disease**

- Apply as supporting for all *TERT* missense variants unless BA1 or BS1 are applicable.

#### **PP3 - Computational evidence shows deleterious effect**

- For missense variants: REVEL >0.55^6,12^.
- Can search variant here: <http://database.liulab.science/dbNSFP> (Make sure to select hg19).

#### **PP4 - Patient's phenotype/family history highly specific for the disease**

- Apply if two or more phenotypic features of TBD.

**PP5 - Sources report as pathogenic** – not applicable

#### **BENIGN CRITERIA**

Where possible criteria were applied as below. As there is a paucity of LB/B variants that have undergone functional testing, variants that met BS1 were considered LB for the purposes of this analysis.

Variants Ala202Thr, Glu280Lys and Pro380Ser have been considered LB given there are a very high number VarSome views and ClinVar LB entries^13,14^. The Pro380Ser variant has also been seen co-segregating with the DKC1 c.961C>A^15^.

#### **BA1 - In population studies >5%**

- Allele frequency is 0.5% or greater in ESP, 1000G, gnomAD or ExAC^1^.

#### **BS1 - High allele frequency**

- Using cardioDB (<http://cardiodb.org/allelefrequencyapp/>)^16^:

DC

- Estimated prevalence of DC: 1:100000^17^.
- *TERT* variants estimated to account for approximately 5% of DC^17^.
- 9/75 patients with DC phenotype and WES performed found to have suspicious *TERT* variant (12%)^18^.
- 17/194 index cases in the UK DC registry (9%)^19^.
- 3/135 Nordic patients with suspected TBD harboured suspicious *TERT* variant (2%)^20^.
- 15/80 patients with inherited and acquired bone marrow failure harboured suspicious *TERT* variant (19%)^21^.
- 7/38 consecutive patients presenting with bone marrow failure or pulmonary fibrosis to Johns Hopkins Hospital from 2005-2009 for genetic evaluation found to have suspicious *TERT* variant (18%)^22^.
- 6/153 probands from The University of Chicago Inherited Hematologic Disorders Registry (4/22 probands in families with AA and IPF)^8^.
- Monoallelic, Prevalence of 1/100000, allelic heterogeneity 0.05, genetic heterogeneity 0.2, penetrance 0.02 = Maximum tolerated reference AC: 0.

AML/MDS

- 2/20 families with suspicion for familial MDS/AML (10%)^23^.
- Estimated lifetime prevalence of AML/MDS: 1:100.
- 41/1514 MDS patients (2.7%) found to have rare *TERT* variant^24^.
- 11/133 consecutive patients with AML harboured a rare germline *TERT* variant (8%)^25^.
- Monoallelic, Prevalence of 1/100, allelic heterogeneity 0.05, genetic heterogeneity 0.05, penetrance 0.02 = Maximum tolerated reference AC: 90, Maximum credible population AF: 0.000625.

IPF

- Prevalence of IPF is 0.57 to 4.51/10000 (Asia Pacific), 0.33 to 2.51/10000 (Europe), and 2.40 to 2.98/10000 (North America)^26^.
- 1:50 patients with IPF have a first degree relative with IPF. Prevalence of familial IPF estimated at 1.34/1,000,000^27^.
- 6/46 IPF families found to have potentially causative *TERT* variant (13%)^28^.
- 5/73 families in the Vanderbilt Familial Pulmonary Fibrosis Registry found to have a potentially causative *TERT* variant (7%)^29^.
- Monoallelic, Prevalence of 1/800,000, allelic heterogeneity 0.05, genetic heterogeneity 0.1, penetrance 0.02 = Maximum tolerated reference AC: 0.

CLD

- Prevalence of cirrhosis: Western Europe approximately 1500/100000, high income North America 375/100000, Oceania 600/100000^30^.
- 9/134 patients with cirrhosis harboured a missense variant in *TERT* (7%)^31^.
- 6/170 patients with non-alcoholic fatty liver disease or hepatocellular carcinoma harboured suspicious *TERT* variants (4%)^32^.
- 4/120 patients with HCC associated with cirrhosis (3%)^33^.
- Monoallelic, Prevalence of 1/100, allelic heterogeneity 0.05, genetic heterogeneity 0.04, penetrance 0.02 = Maximum tolerated reference AC: 74, Maximum credible population AF: 5e-04.

AA

- 5/363 patients with AA phenotype found to have a *TERT* variant (1%)^34^.
- 2/96 Japanese children with acquired AA found to have rare *TERT* variant (2%)^35^.

To determine AF for a given allele review the Popmax Filtering AF calculation in gnomAD. If it is > **0.000625** apply BS1.

**BS2 - Observed in a healthy adult** – not applicable

**BS3 - Functional studies show no damaging effect** – not applicable

**BS4 - Lacking segregation** – not applicable

**BP1 - Does not fit mutational spectrum** – not applicable

**BP2 - Observed with another pathogenic variant** – not applicable

**BP3 - In-frame deletions/insertions in a region without a known function** – not applicable

#### **BP4 - Computational evidence shows no impact**

- For missense variants: REVEL <0.43^6,12^ and spliceAI <0.38.
- Can search variant here: <http://database.liulab.science/dbNSFP> (Make sure to select hg19)
- Apply for synonymous variants alongside BP7 supporting if criteria for BP7 are met (to force to a C2).

**BP5 - Alternate molecular basis for disease** – not applicable

**BP6 - Sources report as benign –** not applicable

#### **BP7 - Predicted as benign synonymous variant**

- Synonymous variants only.
- PhyloP <0.1 (as per RUNX1 ClinGen guidelines)^7^. This number is present in the Alamut summary against nucleotide conservation.
- Check splice predictions (Alamut splice predictors AND Splice AI: <https://spliceailookup.broadinstitute.org/> ).
  - SSF and MES (Alamut) predict either an increase in the canonical splice site score or a decrease in the canonical splice site score by **no more** than 10%, and no putative cryptic splice sites are created.
  - No recommended Splice AI score. Arbitrary cut-off of 0.75 recommended - if >0.75 this criterion should not be applied. Must be <0.75.
- Also applicable to intronic and UTR variants detected.

#### ***TERC***

#### **PATHOGENIC CRITERIA**

The following application of ACMG criteria was applied.

#### **PVS1 - Null variants-** not applicable

#### **PS1 - Established Pathogenic Variant-** not applicable

#### **PS2 - Confirmed de novo variant**

- Also see PM6.
- Apply as usual, but likely to be rarely applicable due to age of onset of phenotype. Paternity confirmation by SNP array or SNP assessment of NGS sample (needs to be specifically stated).
- See: <https://clinicalgenome.org/site/assets/files/3461/svi_proposal_for_de_novo_criteria_v1_1.pdf>

Points per de novo occurrence where paternity is confirmed:

| **Phenotypic consistency** | **Points per proband (mutually exclusive selection)** |
| --- | --- |
| Two or more phenotypic features of a TBD | 2 |
| One phenotypic feature of TBD | 0.5 |

| **PS2_Supporting** | **PS2_moderate** | **PS2_strong** | **PS2_VeryStrong** |
| --- | --- | --- | --- |
| 0.5 | 1 | 2 | 4 |

#### **PS3 - Functional studies support damaging effect** – not applicable as under study

#### **PS4 - Enrichment in cases**

See *TERT* sensitivity analysis comments.

If the variant is absent or present once in gnomAD^10^ and there is evidence of at least two reports of the variant occurring in a case (case defined as a ClinVar^13^ entry, UniProt^36^ entry, report in the literature*, three or more views in Varsome^14^) this was deemed sufficient evidence for application of PS4. More than 10 views in Varsome^14^ was also considered as at least two reports of the variant occurring in cases.

PS4_moderate: single report in the literature OR ClinVar^13^ OR UniProt^36^ AND Varsome^14^ views were between two-three

PS4_supporting: single report in the literature OR ClinVar^13^ OR UniProt^36^ AND Varsome^14^ views were less than two

*If only single case report AND single ClinVar/Uniprot^13,36^ entry this was counted as one case given that there is a high likelihood that a single case would be published and submitted to ClinVar^13^.

#### **PM1 - Mutational hotspot/functional domain-** not applicable

#### **PM2 - Absent from population studies**

- Supporting level only. Zero times in gnomAD^10^.

**PM3 - Detected in trans** – not applicable

#### **PM4 - Protein length changes-** not applicable

#### **PM5 - Novel missense change-** not applicable

#### **PM6 - Assumed de novo**

- Also see PS2.
- Points per de novo occurrence where paternity is unconfirmed:

| **Phenotypic consistency** | **Points per proband (mutually exclusive selection)** |
| --- | --- |
| Two or more phenotypic features of a TBD | 1 |
| One phenotypic feature of TBD | 0.25 |

| **PM6_Supporting** | **PM6_moderate** | **PM6_strong** | **PM6_VeryStrong** |
| --- | --- | --- | --- |
| 0.5 | 1 | 2 | 4 |

#### **PP1 - Co-segregation with disease**

|  | **PP1 strong** | **PP1 moderate** | **PP1 supporting** |
| --- | --- | --- | --- |
| Phenotype | One or more phenotypic features of a TBD | One or more phenotypic features of a TBD | One or more phenotypic features of a TBD |
| Number of meioses | 7 or more | 5-6 | 3-4 |

- Enumeration based on literature search (google and mastermind^11^) and inhouse database.
- Meioses within and between families weighted the same.
- Only include genotype positive and phenotype positive meioses.
- Do not include the proband/s in the count.
- Cases of parent-child only literature reports: this can be considered as 1 meiosis.
- See PP1_BS4 presentation for more details.

#### **PP2 - Missense variants are a common mechanism of disease-** not applicable

#### **PP3 - Computational evidence shows deleterious effect-** not applicable

#### **PP4 - Patient's phentoype/family history highly specific for the disease**

- Apply if two or more phenotypic features of TBD.

**PP5 - Sources report as pathogenic** – not applicable

#### **BENIGN CRITERIA**

Where possible criteria were applied as below. As there is a paucity of LB/B variants that have undergone functional testing, variants that met BS1 were considered LB for the purposes of this analysis.

#### **BA1 - In population studies >5%**

- Allele frequency is 0.5% or greater in ESP, 1000G, gnomAD or ExAC^1^.

#### **BS1 - High allele frequency**

- Using cardioDB^16^ (<http://cardiodb.org/allelefrequencyapp/>):

DC

- Estimated prevalence of DC: 1:100000^17^
- TERC variants estimated to account for approximately 5% of DC^17^.
- 30/194 index cases in the UK DC registry (15%)^19^.
- 9/338 index cases in the UK DC registry (new cohort) (3%)^18^.
- 4/135 Nordic patients with suspected TBD harboured suspicious *TERC* variant (3%)^20^.
- 3/38 consecutive patients presenting with bone marrow failure or pulmonary fibrosis to Johns Hopkins Hospital from 2005-2009 for genetic evaluation found to have suspicious *TERC* variant (8%)^22^.
- 4/153 probands from The University of Chicago Inherited Hematologic Disorders Registry (2/22 probands in families with AA and IPF)^8^.
- Monoallelic, Prevalence of 1/100000, allelic heterogeneity 0.05, genetic heterogeneity 0.2, penetrance 0.02 = Maximum tolerated reference AC: 0.

AML/MDS

- Estimated lifetime prevalence of AML/MDS: 1:100.
- 0/133 consecutive patients with AML harboured a rare germline *TERC* variant (0%)^25^.

IPF

- Prevalence of IPF is 0.57 to 4.51/10000 (Asia Pacific), 0.33 to 2.51/10000 (Europe), and 2.40 to 2.98/10000 (North America)^26^.
- 1:50 patients with IPF have a first degree relative with IPF. Prevalence of familial IPF estimated at 1.34/1,000,000^27^.
- 1/46 IPF families found to have potentially causative *TERC* variant (2%)^28^.
- 1/73 families in the Vanderbilt Familial Pulmonary Fibrosis Registry found to have a potentially causative *TERC* variant (1%)^29^.
- Monoallelic, Prevalence of 1/800,000, allelic heterogeneity 0.05, genetic heterogeneity 0.1, penetrance 0.02 = Maximum tolerated reference AC: 0.

CLD

- Prevalence of cirrhosis: Western Europe approximately 1500/100000, high income North America 375/100000, Oceania 600/100000^30^.
- 1/134 patients with cirrhosis harboured a missense variant in *TERC* (1%)^31^.
- 0/120 patients with HCC associated with cirrhosis (0%)^33^.
- Monoallelic, Prevalence of 1/100, allelic heterogeneity 0.05, genetic heterogeneity 0.01, penetrance 0.01 = Maximum tolerated reference AC: 40, Maximum credible population AF: 0.00025

AA

- 0/96 Japanese children with acquired AA found to have rare *TERC* variant (0%)^35^.

To determine AF for a given allele review the Popmax Filtering AF calculation in gnomAD. If it is **> 0.00025** apply BS1.

**BS2 - Observed in a healthy adult** – not applicable

**BS3 - Functional studies show no damaging effect** – not applicable

**BS4 - Lacking segregation** – not applicable

**BP1 - Does not fit mutational spectrum** – not applicable

**BP2 - Observed with another pathogenic variant** – not applicable

**BP3 - In-frame deletions/insertions in a region without a known function** – not applicable

#### **BP4 - Computational evidence shows no impact**– not applicable

**BP5 - Alternate molecular basis for disease** – not applicable

**BP6 - Sources report as benign –** not applicable

#### **BP7 - Predicted as benign synonymous variant-** not applicable

### OddsPath calculations

#### DTA

16 variants (12 LP/P and 4 LB/B) with DTA results were identified. A functionally abnormal result was considered as less than 75% of wild type activity. 11 functionally abnormal (all true positive) results and 5 functionally abnormal (4 true negative and 1 false positive) results were identified (Table 1). These results were then entered into the Bayesian calculator.

#### TRAP

20 variants (15 LP/P and 5 LB/B) with TRAP results were identified. The Pro380Ser (LB) and Arg951Trp (LP) were reported as associated with 80% of wildtype activity. In a perfect binary analysis 80% activity was considered functionally abnormal. This resulted in 16 functionally abnormal (13 true positive and 3 false positive) results and 4 functionally normal (2 true negative and 2 false negative) results. This increased number of false negative and false positive results appeared to falsely improve the OddsPath calculation (Table S1). This is demonstrated by serial removal of the false negative and false positive results. Omission of false negative results was associated with a significant reduction in the OddsPath score for benign from 0.067 to 0.208 (Table S2). Omission of false positive results significantly reduced the OddsPath score for pathogenic from 5.0 to 1.6 (Table S3).

Table S1

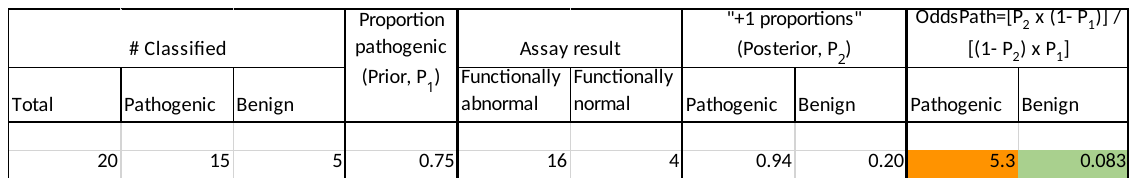

Table S2

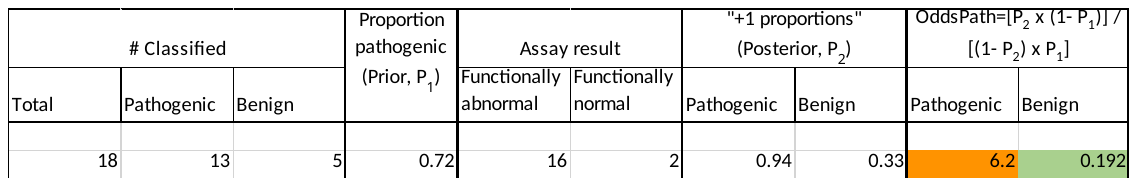

Table S3

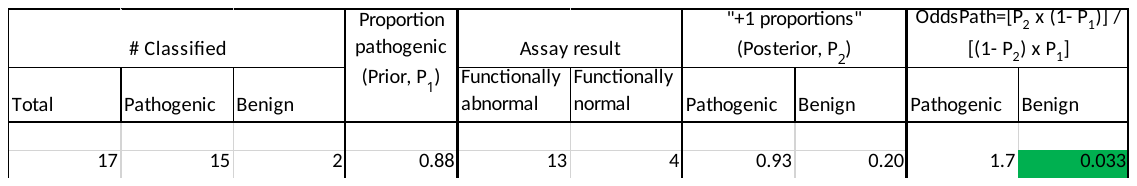

Using an alternative calculation where the results associated with the variants Pro380Ser and Arg951Trp were considered as indeterminant resulted in 14 functionally abnormal (12 true positive and 2 false positive) results and 4 functionally normal (2 true negative and 2 false negative) results. The OddsPath calculations for pathogenic and benign were 4.7 and 0.083 respectively (Table S4).

Table S4

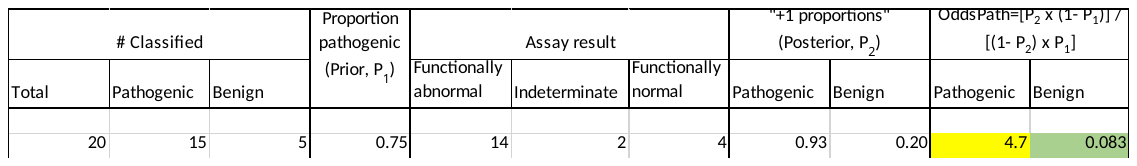

Given the impact of false positive and false negative results a range for the OddsPath score was provided (supplementary results S7).

### Sensitivity analysis *TERT*

Relaxed PS4

PS4 is very challenging to apply for TBD related genes for multiple reasons.

1. The associated phenotype to define a ‘case’ is not defined. It is likely that the phenotypic consequences of *TERT* and *TERC* variants are on a continuous spectrum depending on the functional impact of the variant, anticipation, coinherited genetic modifiers and environmental exposures.
2. Published cohort studies often involve selected cohorts of cases, increasing the risk of ascertainment bias.

The largest pooled cohort of unselected cases is in Euro/North American AML/MDS cases (2393 alleles)^23-25^. On reviewing confidence intervals for OR calculated using case cohort data and gnomAD^10^ as the control data, 3 probands would likely be significant even if a relativity poorly represented Ethnicity was reviewed (e.g. East Asian) if the allele frequency was less than 4 in gnomAD^10^ (Table S5).

Table S5

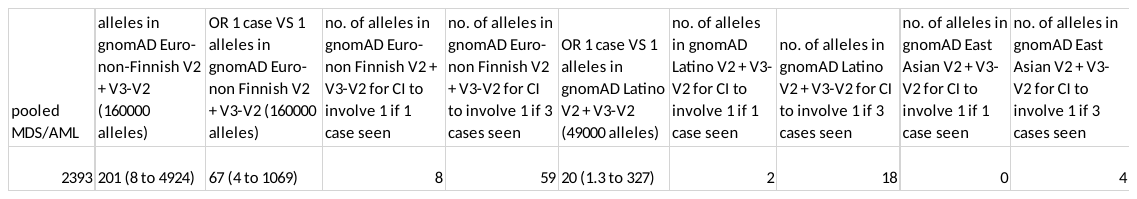

MDS (myelodysplastic syndrome), AML (acute myeloid leukaemia), OR (odds ratio), CI (confidence interval)

In light of this, if the variant is absent or present once in gnomAD^10^ and there is evidence of at least two reports of the variant occurring in a case (case defined as a ClinVar^13^ entry, UniProt^36^ entry, report in the literature*, three or more views in Varsome^14^) this was deemed sufficient evidence for application of PS4. More than 10 views in Varsome^14^ was also considered as at least two reports of the variant occurring in cases.

PS4_moderate: single report in the literature OR ClinVar^13^ OR UniProt^36^ AND Varsome^14^ views were between two-three.

PS4_supporting: single report in the literature OR ClinVar^13^ OR UniProt^36^ AND Varsome^14^ views were less than two.

*If only single case report AND single ClinVar/Uniprot^13,36^ entry this was counted as one case given that there is a high likelihood that a single case would be published and submitted to ClinVar^13^.

Relaxed PM1

The functional domains of *TERT* show significant constraint to missense variants, however the domains are large and there are no clear hotspot residues. Considering somatic data for *TERT* is also problematic as somatic variants are most commonly associated with gain of function. PM1 was applied at a very low strength level o ensure only the most well-established LP/P variants were used to assess functional genomic data,. To review the impact of applying PM1 at higher strength multiple alternative PM1 rules were applied as follows:

1. PM1_supporting: In a functional domain and 1 or more somatic mutations reported at codon.

PM1_moderate: In the TRBD domain.

2. PM1_moderate: In a functional domain.
